# Features of creatine-kinase in COVID-19 patients within various specific periods: A cohort study

**DOI:** 10.1101/2020.10.28.20221119

**Authors:** Shanshan Wan, Gaojing Qu, Hui Yu, Haoming Zhu, Guoxin Huang, Lei Chen, Meiling Zhang, Jiangtao Liu, Bin Pei

**Author notes:** Corresponding authors: Professor Jiangtao Liu, Department of Orthopedics, Xiangyang No.1 People’s Hospital, Hubei University of Medicine, Xiangyang 441000, Hubei Province, China. Professor Bin Pei, Center of Evidence-based Medicine, Xiangyang No.1 People’s Hospital, Hubei University of Medicine, Xiangyang 441000, Hubei Province, China. **Declaration of interests:** We declare no competing interests.

## Abstract

**Background:** Coronavirus disease 2019 (COVID-19) has been declared as a threat to the global. Due to the lack of efficient treatments, indicators were urgently needed during the evolvement of disease to analyze the illness and prognosis, and prevent the aggravation of COVID-19.

**Methods:** Patients’ general information, clinical type, all CK values and outcome were collected. CK value of all cases during disease course started from different initial time were analyzed.

**Results:** All cases underwent 504 tests of CK since symptom onset and the median value was 51.7 (35.0-91.5) U/L. The first median value on the day 8 from exposure onset was 78.1 (69.1-85.8) U/L then showed an upward trend from the day 8 to the day 12 (reaching a peak of 279.3 U/L), finally showed a fluctuation decline after the day 12. The CK median value in critical cases reached the peak (625.5 U/L) on the transforming date, and then decreased rapidly to the normal range. Before death, the CK median value in dead cases firstly increased until the day −14 with a peak as 470.0 U/L, then decreased with fluctuation until day −2, and finally increased again on the day 0.

**Conclusions:** CK reached its peak on the day when it became critical type, dynamic detection of CK can guide clinical judgment of prognosis. The increase of CK is a high risk factor of death. Severe cell damage 2 weeks before death might determines the outcome of the disease even if CK drops to the normal range afterward.

## 1. Introduction

In 2019, coronavirus disease (COVID-19) was caused by severe acute respiratory syndrome coronavirus type 2 (SARS-CoV-2), which has spread around the world[1-3]. Creatine Kinase (CK) usually exists in the cytoplasm and mitochondria of the heart, skeletal muscle and brain tissues of animals. It is an important kinase directly related to intracellular energy transport, muscle contraction and ATP regeneration. It can reversibly catalyze the following reactions: ATP + creatine→ADP + phosphocreatine[4, 5]. CK is abundant in ATP rapidly regenerated cells and plays an important role in ion transmembrane transport, phagocytosis of phagocytes, regulation of glycolysis and release of substances dependent on ATP neurotransmission in brain synapses[6].

In 1986, Jaffe *et al*. reported that the CK increased significantly in the early stage of acute myocardial infarction[7]. Chen *et al*. indicated that myocardial zymogram was abnormal in most COVID-19 patients, and CK increased in 13 cases (13%)[8]. Two Meta analysis showed that patients with elevated CK levels had a significantly increased risk of serious illness or admission to ICU, and there was significant difference between severe and non-severe COVID-19 patients (P< 0.01)[9, 10]. As reported by retrospective analysis, the level of CK in the severe group was remarkably higher than those in the non-severe group, and the increase of CK could be used as a predictor of the severity[11, 12]. Chen Tao *et al*. reported the clinical manifestations of the fatal cases in COVID-19 patients and indicated that the level of CK in the death group was significantly higher than that in the rehabilitation group[13]. A univariate analysis showed that CK > 185 U/L was associated with the death of COVID-19 patients[14].

Overall, there lack of systematic research on the changes of CK with different initial time or during transformed period of severity classification, and the relationship between CK and the evolving illness of COVID-19 cases. In order to reveal the features of CK during different periods, we included all confirmed COVID-19 cases hospitalized in Xiangyang No.1 People’s hospital, and analyzed all CK results tested in outpatient and hospitalization.

## 2 Methods

### 2.1 Study design

This study was a bidirectional observational cohort study and the cohort was established on Feb 9, 2020. All suspected and confirmed cases of COVID-19 admitted to the Xiangyang No.1 People’s Hospital Affiliated Hospital of Hubei University of Medicine according to the Diagnosis and Treatment Protocol for Novel Coronavirus Pneumonia (1^st^-7^th^ editions) were included in this study[15]. The retrospective data were traced back to Jan 22, 2020 and the follow-up was carried out until Mar 28, 2020.

### 2.2 Patients

Patients were classified into mild, moderate, severe, and critical type according to diagnosis and treatment protocol[15]. The classification results were cross-checked by two experts, and a third specialist involved if the inconsistency existed. The study was approved by the ethics review board of Xiangyang No.1 People’s Hospital (No. 2020GCP012) and registered at the Chinese Clinical Trial Registry as ChiCTR2000031088. Informed consent from patients has been exempted since this study does not involve patients’ personal privacy neither incur greater than the minimal risk.

### 2.3 Data collection

Data were extracted from the hospital information system by two groups and then cross-checked. Sex, age, CK value, exposure date, disease onset date, transformed date of clinical classification on severity type, outcome, death date were collected.

### 2.4 Outcome measures

In this cohort, symptom onset was regarded as disease onset and the corresponding date was set as the “day 1” to record data from symptom onset. For patients with clear exposure date, the exposure date was set as the first day to record course when the CK changes from exposure onset. The day when the clinical type changes to severe or critical type was regarded as the “day 0” to study the distribution of CK before and after the type transformation. Death date was set as the “day 0” when studying the CK situation during the days before death. Since the time from symptom onset to discharge was 26.9±9.2 days, we recorded and statistically analyzed the data from day 1 to day 30. The distributions of CK median value in course/period were plotted against the time unit of 1 day, 2 days, and 5 days (T1, T2, T3…Tn represented the 5-days unit successively).

### 2.5 Statistical analysis

All statistical analyses were performed through SPSS 20.0. Binary data were described by frequency and percentage. The normality of continuous data was checked. Mean and standard deviation were used to describe variables with normal distribution; otherwise, median (interquartile, IQR) was applied. Categorical data were described as frequency (%); the chi-square test was applied to assess significance between groups. All graphs were processed through GraphPad Prism 8.0 and Photoshop CC 14.2 software.

## 3. Results

This studying included all of the suspected and laboratory-confirmed 542 patients till Feb 28, 2020. Among the 542 cases, the nucleic acid tests in 142 cases were positive. Excluding 2 infants, and 9 cases that transferred from other hospital whose data cannot be traced, 131 cases were included in the further study finally.

### 3.1 General information

Among the included 131 cases, there were 63 males and 68 females, and the average age was 50.1±17.1 years old. The average time from exposure to symptom onset was 9.9±5.3 days, from symptom onset to admission was 4.5±3.1 days, from symptom onset to discharge was 26.9± 9.2 days, from symptom onset to severe type was 8.2±5.3 days, from symptom onset to critical type was 11.8±5.2 days, from severe to critical type was 4.3±5.8 days, from symptom onset to death was 18.4 ±9.8 days, and the length of hospitalization was 22.4± 8.7 days. The 121 cases were discharged from the hospital while 10 cases died.

### 3.2 CK test results

The 131 cases underwent 37 laboratory indicators contained 24052 tests in outpatient and hospitalization. This cohort included all of the CK results, totally 504 times of test, account for 2.1% of the all results of indicators. The normal range of CK value is 50.0-310.0 U/L.

### 3.3 The features of CK from symptom onset

The 131 cases underwent 504 tests of CK from the first day after symptom onset to the day 30, the median value was 51.7 (35.0-91.5) U/L. 5.6% of the tests were higher than the upper limit of normal value (ULN). In T1-T6, the CK median value was 81.5 (57.1-134.0) U/L, 72.7 (43.6-160.5) U/L, 49.6 (37.0-75.5) U/L, 46.1 (29.9-68.0) U/L, 43.0 (32.5-60.3) U/L, and 37.2 (26.6-51.9) U/L, respectively; and the abnormal rate was 9.0%, 10.6%, 4.4%, 4.2%, 1.3%, and 1.6%, respectively (Table 1). According to the changing trend plot per day, the CK value decreased during day 1 (97.0 U/L) to day 2 (88.0 U/L), increased during day 2 to day 4 (101.1 U/L, the peak value), then decreased with fluctuation after day 4 (Fig. 1A); according to the changing trend plot per 5 days, the CK value decreased during T1-T6, and the peak value (81.5 U/L) distributed at T1. No CK median values exceed the normal range (Fig. 1B).

**Table 1.** Median value and abnormal percentage of CK from symptom onset in 131 patients

| Period | Median value (U/L) | Total tests | Abnormal Percentage (%) |
| --- | --- | --- | --- |
| Total course | 51.7 (35.0-91.5) | 504 | 5.6% |
| T1 | 81.5 (57.1-134.0) | 67 | 9.0% |
| T2 | 72.7 (43.6-160.5) | 113 | 10.6% |
| T3 | 49.6 (37.0-75.5) | 90 | 4.4% |
| T4 | 46.1 (29.9-68.0) | 96 | 4.2% |
| T5 | 43.0 (32.5-60.3) | 76 | 1.3% |
| T6 | 37.2 (26.6-51.9) | 64 | 1.6% |

**Fig. 1.**
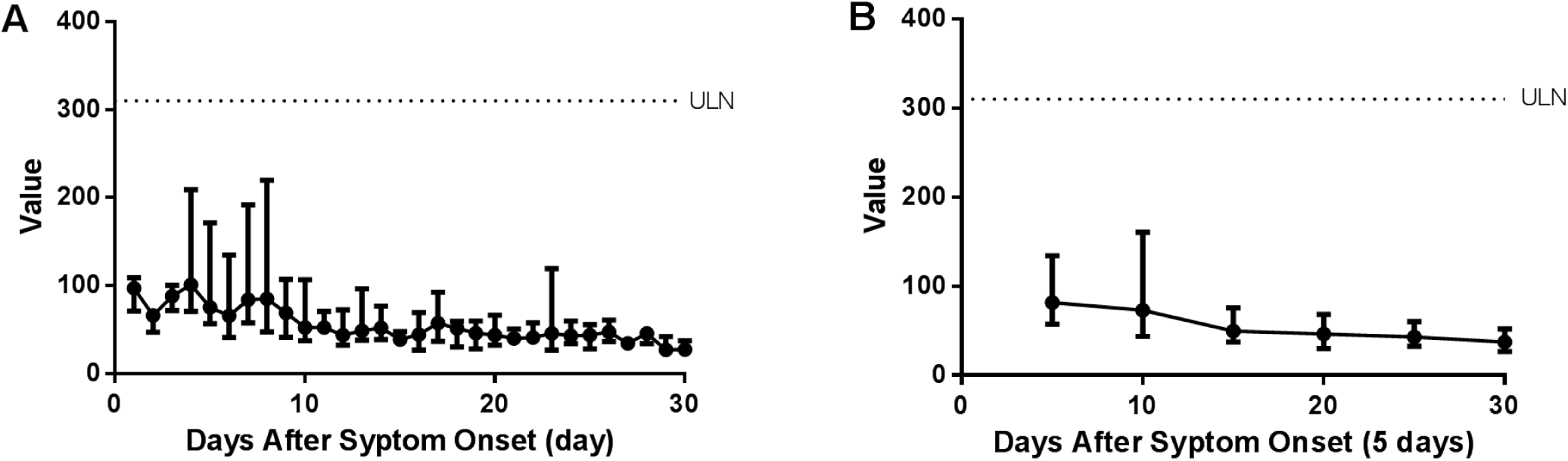
The distribution over time of CK median in total cohort from symptom onset per day (A) and per 5 days (B).

### 3.4 The features of CK from exposure onset

We followed up all of the cases and finally defined 42 cases with exact exposed day. The 42 cases underwent 207 tests and the median was 57.2 (38.0-124.6) U/L, among which 9.7% of the tests were higher than the ULN. The changes of CK median from exposure onset indicated that the first median value was 78.1 (69.1-85.8) U/L on the day 8, the CK value increased during day 8 to day 12 (279.3 U/L, the peak value), then decreased with fluctuation after day 12. No CK median values exceed the normal range (Fig. 2A).

**Fig. 2.**
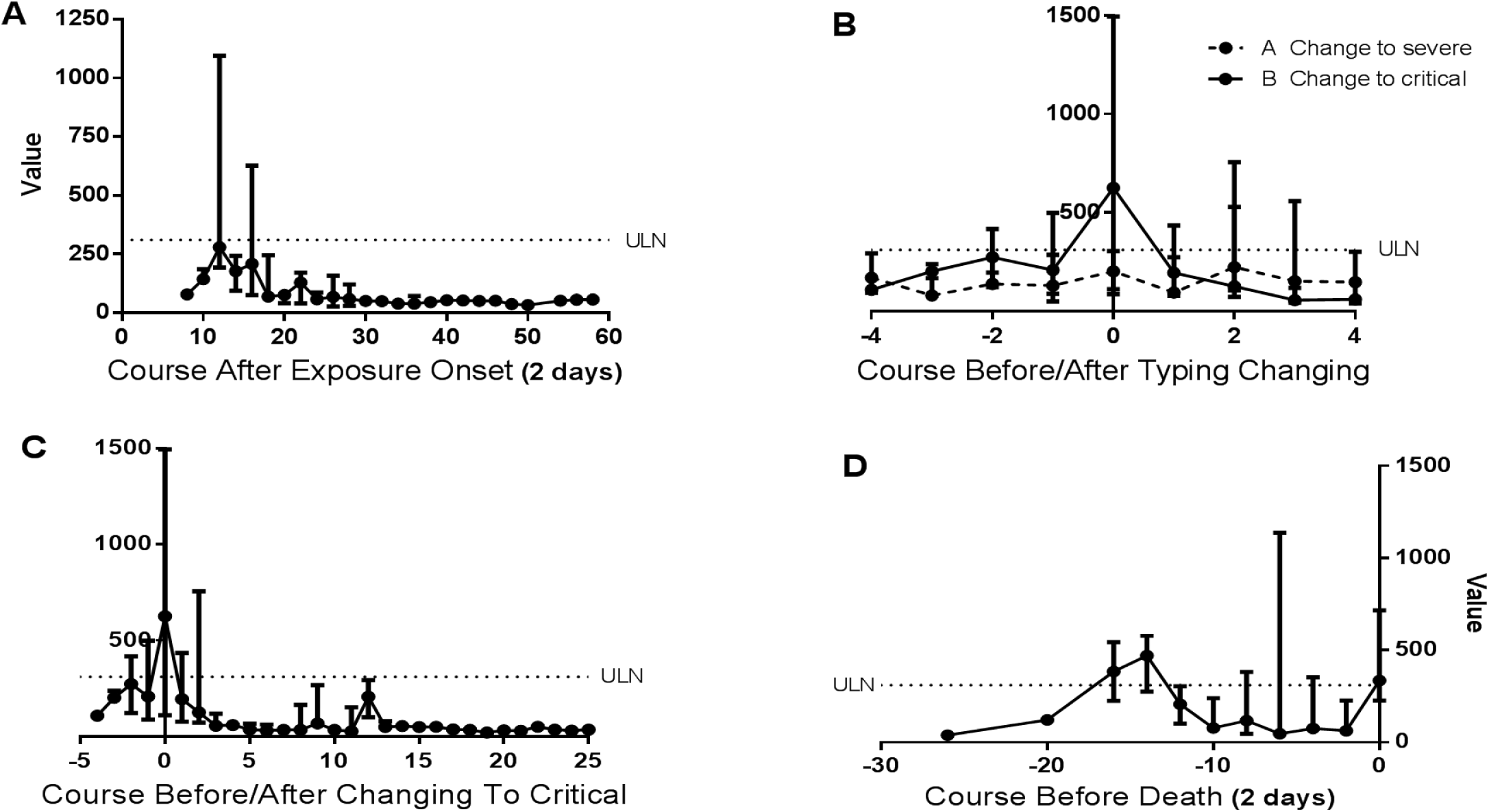
The distribution of CK median from exposure onset (A), before and after transformation into severe type (B) and critical type (B, C), and before death (D).

### 3.5 The features of CK before and after transformation into severe and critical type

The transformed date developed into severe type was set as “0” in 21 severe cases and 18 critical cases, the changes that 4 days before and after “0” was plotted. Above the 74 test results of severe cases, the median was 141.6 (59.6-291.2) U/L, and 23.0 % of them were higher than the ULN. The tendency showed that the CK value fluctuate between 100.0 U/L and 200.0 U/L, and the peak value (222.73 U/L) distributed at day +2, all CK median values are within the normal range. Similarly, changing trend 4 days before and after transforming date of developing into critical type in 18 cases was plotted. Above the 50 test results, the median was 142.7 (68.4-366.9) U/L, and 30.0% of them were higher than the ULN. The chart showed that the CK value was increased during day −4 (107.7 U/L) to day 0 (625.5 U/L, the peak value) while decreased during day 0 to day +4 (60.0 U/L). The abnormal value was found on the day 0 (Fig. 2B).

### 3.6 The features of CK before died

The 55 tests were conducted between the day −26 to day 0 with the abnormal rate as 30.9 % and the median value as 117.5 (51.0-371.6) U/L in the 10 dead cases. The tendency showed that the CK value increased during day −26 (38.8 U/L) to the day −14 (470.0 U/L, the peak value), then decreased with fluctuation during day −14 to day −2 (63.2 U/L), finally increased during day −2 to day 0 (336.1 U/L). The abnormal period was day −16 to day −14 and day 0 (Fig. 2D).

## 4. Discussion

When studying the course from symptom onset, the median of CK was 51.7 (35.0-91.5) U/L, and 5.6% of the total tests exceeded ULN. The peak value of CK appeared on the early stage of disease (day 4 and T1 in the changes per day and per 5 days, respectively), suggesting that CK might have potential value for the diagnosis at the early stage. No CK median values exceed the normal range, which shows that for COVID-19 patients, the increase of CK is not beyond the upper limit of normal value. Further research were in need in the future work. Symptom onset always regarded as the disease onset, while virus replication in the infected host induced tissue damage and inflammatory response before symptom onset. Symptom onset once the viral load and/or damage reached a certain extent, thus the actual whole course should be recorded from exposure onset. As shown in Figure 2A, the first test appeared on the day 8 since exposure onset, then increased to a peak (279.3 U/L) on the day 12. About 12 days after exposure was an important time period for CK detection, the CK value of this period is helpful for us to judge the severity and prognosis of COVID-19. The peak value appeared early, indicated that early monitoring of CK value contribute to better evaluate the disease and is of great significance to the graded treatment of patients, which is similar to the results reported by Guzik *et al* [16].

Due to the differences of disease progression, the time for patients to transform from moderate type to severe and critical type was quite different. No significant increase in CK was found during the transformation from moderate type to severe type, but CK increased and reached its peak on the day when severe type transformed to critical type. According to the results, CK increased during day −4 to day 0, decreased during day 0 to day 4; the peak value (625.5 U/L) distributed at day 0 (the day of transformation to critical type), which was higher than the ULN. In order to further observe the change, we extended the change trend chart (Fig. 2C). It can be seen that after reaching the peak on the day it became critical type (day 0), the CK median value quickly returned to the normal range on the day 1 (194.6 U/L), the maintenance time is short (1 day), indicating that serious tissue damage occurred before/after the transition day, which might reflect the degree of tissue injury of COVID-19, and the more serious the injury, the more serious the disease and the prognosis, this conclusion is consistent with a Meta analysis reported by Zhang *et al*[17]. Thus dynamic detection of CK value may be helpful to monitor the severity of COVID-19 patients.

The time from symptom onset to death was 18.3±9.65 days, which was relatively dispersed. Thus, it could not describe the features of CK before death exactly. In our study, we made a CK time distribution diagram with the date of death as “day 0”. As shown in the Fig. 2D, the CK median value reached the peak on the day −14, which reveals that tissue damage might be heaviest on the day −14. After the day −12, it gradually decreased to the normal range, which was similar to the transition of critical type. This indicates that severe cell damage 2 weeks before death might determines the outcome of the disease even if CK drops to the normal range thereafter. This shows that the increase of CK beyond the ULN is a risk factor of death, and early dynamic detection of CK value contribute to the analysis and evaluation of prognosis in COVID-19 patients.

## 5. Limitation

The current study has several limitations. First, the cases included in our cohort were enrolled from single hospital, the evidence value was limited. Second, due to the limited sample size, the data could not be analyzed and described daily, and some information has been lost.

## 6. Conclusion

Early monitoring of CK value is helpful to better evaluate the COVID-19. The CK value reached its peak on the day when it became critical type, severe cell damage 2 weeks before death might determines the outcome of the COVID-19, early dynamic detection of CK value is of great help to analyze and judge the changes of the illness.

## Data Availability

Anyone who wishes to obtain the original data of this study with reasonable purposes can contact the correspondent author via email.

